# Deep immune profiling of chronic rhinosinusitis in allergic and non-allergic cohorts using mass cytometry

**DOI:** 10.1101/2023.12.21.23300286

**Authors:** Fana Alem Kidane, Lena Müller, Marianne Rocha-Hasler, Aldine Tu, Victoria Stanek, Nicholas Campion, Tina Bartosik, Mohammed Zghaebi, Slagjana Stoshikj, Daniela Gompelmann, Andreas Spittler, Marco Idzko, Sven Schneider, Julia Eckl-Dorna

## Abstract

**Background:** Chronic rhinosinusitis (CRS) is characterized by persistent nasal and paranasal sinus mucosa inflammation. It comprises two phenotypes, namely CRS with nasal polyps (CRSwNP) and without (CRSsNP). CRSwNP can be associated with asthma and hypersensitivity to non-steroidal anti-inflammatory drugs (NSAIDs) in a syndrome known as NSAID-exacerbated respiratory disease (N-ERD). Furthermore, CRS frequently intertwines with respiratory allergies.

**Objective:** This study investigated the phenotypic characteristics of peripheral blood mononuclear cells (PBMCs) within cohorts of CRS patients, additionally examining the influence of comorbid respiratory allergies on these parameters.

**Methods:** 24 participants were grouped into controls, CRSsNP, CRSwNP, and N-ERD (*n=6*/group), with half of the patients in each group having respiratory allergies. Levels of cytokines were quantified in nasal secretions and sera. The abundance and phenotypic features of immune cells in PBMCs were evaluated through mass cytometry and clustering methods.

**Results:** N-ERD patients showed heightened type 2 nasal cytokine levels. Mass cytometry analysis revealed increased activated naive B cell levels in CRSwNP and N-ERD, while resting naive B cells were higher in CRSsNP. Th2a cell levels did not differ between CRS subtypes but were significantly elevated in allergic subjects. In CRSwNP and N-ERD patients, naive B cells had a lower CXCR5 and higher CD45RA expression, while NK cells displayed reduced CD56 levels.

**Conclusions:** There are distinct immunological features in PBMCs of CRS phenotypes and allergy, characterized by elevated resting naive B cells in CRSsNP, increased activated naive B cells in CRSwNP and N-ERD, and higher Th2a cell levels in allergic subjects.

**Capsule summary:** This study examines immunological profiles in different phenotypes of CRS with and without comorbid allergy patients, highlighting immune cell intricacies in CRS subtypes and immune differences in CRS and respiratory allergy.

## 1. INTRODUCTION

Chronic rhinosinusitis (CRS) is a complex disease characterized by persistent inflammation of nasal and paranasal sinus mucosa affecting 8-12% of the population ^1^. Clinically, CRS is classified as CRS with nasal polyps (CRSwNP) or without nasal polyps (CRSsNP), with up to 15% of CRSwNP patients also demonstrating comorbid hypersensitivity to aspirin and other nonsteroidal anti-inflammatory drugs (NSAIDs) as well as asthma in a syndrome called NSAIDs-exacerbated respiratory disease (N-ERD)^2^. CRS is further classified into different endotypes based on inflammatory cell infiltrations and cytokine secretions in nasal tissues ^3^. Of these endotypes, the type 2 endotype is predominantly observed in CRSwNP and N-ERD conditions in the Western world ^2,4–6^. Respiratory allergy, another hallmark type 2 disease, affects up to 30% of the population ^7^.CRS and respiratory allergy involve inflammation in the respiratory system and elevated IgE production, but the link between these two conditions is not yet clear ^8^.

Numerous previous studies aiming to characterize various immune cell types in CRS and allergy ^9–18^ employed flow cytometry or immunohistochemistry-based techniques, albeit with limitations on the number of parameters that could be studied simultaneously. Although advancements in single-cell parametrization, driven by novel fluorophores and laser systems, have expanded our understanding of immune cell sub-classes and functional states, spectral overlap in fluorescence-based cytometry often renders comprehensive immune state analysis difficult ^19^.

Mass cytometry, or cytometry by Time-Of-Flight (CyTOF®), offers a potential solution by using heavy-metal isotopes instead of fluorophores, enabling precise quantification of target expression with minimal signal overlap and providing a detailed snapshot of the immune state ^19^. Thus, we used this technique to stain 38 immune cell surface markers simultaneously for in-depth immune profiling of peripheral blood mononuclear cells (PBMCs) in CRS patients. This study’s primary objective was to analyse alterations in abundance and phenotypic characteristics of PBMCs in cohorts of CRS patients and disease controls. Importantly, each group contained an equal representation of patients with respiratory allergies, allowing us to distinguish whether the changes were exclusively linked to CRS or could be influenced by the allergic predisposition.

## 2. MATERIAL AND METHODS

### 2.1. Study subjects and sample collection

Samples (Serum, nasal secretions, PBMCs) used in this study were taken from the CRS biobanks of the Department of Otorhinolaryngology, Medical University of Vienna, with the approval of the Ethical Committee of the Medical University of Vienna (EK Nr. 1492/2023). The biobanks also contained detailed clinical characteristics of the patients as shown in Table I and Supplementary Table E1.

**Table I:** Patient characteristics of study cohorts.

| Group * | Control | CRSsNP | CRSwNP | N-ERD | *P* value |
| --- | --- | --- | --- | --- | --- |
| No of subjects | 6 | 6 | 6 | 6 | n.a |
| Sex, M/F | 2/4 | 4/2 | 4/2 | 5/1 | n.a |
| Median age in years (range) | 40 (24)<sup>+</sup> | 38 (33) | 49 (34) | 54 (22) | 0.0788 |
| Median TPS<sup>**</sup> (range) | 0 (0) | 0 (0) | 4 (6) | 5.5 (5) | 0.0002 |
| Median SNOT-20<sup>***</sup> (range) | 1 (14)<sup>++</sup> | 13 (29)<sup>+++</sup> | 32 (32) | 19 (35) | 0.0023 |

\*Chronic Rhinosinusitis (CRS) without nasal polyps (CRSsNP), CRS with nasal polyps(CRSwNP), Nonsteroidal anti-inflammatory drugs (NSAIDs)-Exacerbated Respiratory Disease (N-ERD); \*\*Total Polyp Score (TPS); \*\*\* 20-item Sinonasal Out Come Test (SNOT-20); <sup>+</sup> datum from one participant was missing; <sup>++</sup> datum from one participant was missing, <sup>+++</sup> data from two participants were missing; n.a, not applicable

### 2.2. Sample preparation

Sera, nasal secretions, and PBMCs were prepared following established procedures and are outlined in detail in the online repository.

### 2.3. IgE measurement

Allergen-specific IgE levels were measured in serum using the ALEX® Allergy Explorer assay (Allergy Explorer version 2, MacroArray Diagnostics, Vienna, Austria), encompassing over 280 allergen extracts and molecular allergens according to the manufacturer’s instructions, which is briefly summarized in the online repository. A description of the main respiratory allergens used to determine a patient’s allergic status in Fig. 1 of this manuscript is also provided in Supplementary Table E2.

**Figure 1:**
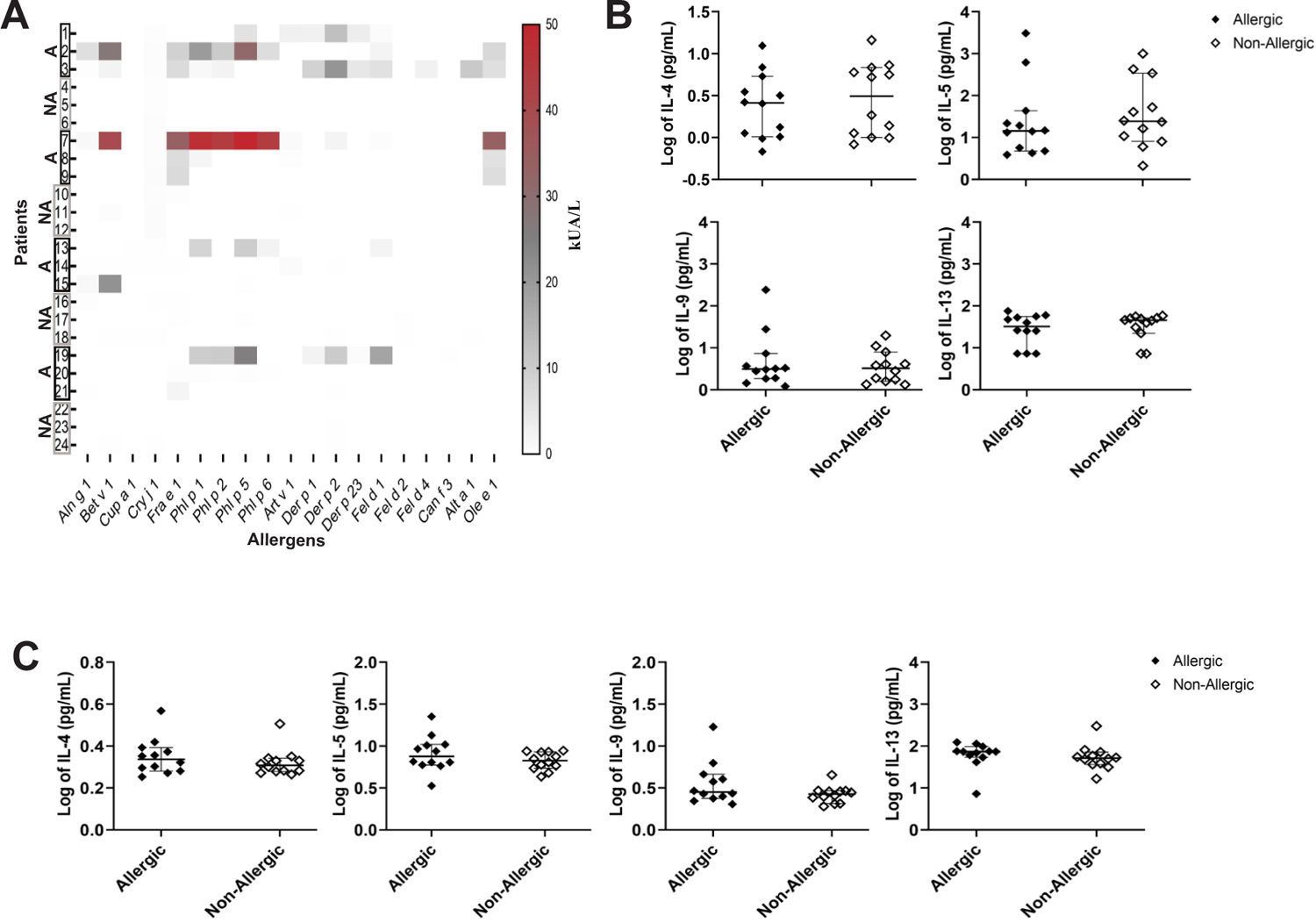
Allergic sensitization, nasal and serum type 2 cytokine levels in patient cohorts. (A) Heat map representing specific IgE levels to selected respiratory allergens in sera of allergic (A, *n*=12) and non-allergic subjects (NA, *n*=12). The color gradient from white to red represents increasing quantities of allergen-specific IgE antibodies (kUA/L). (B, C) Log values of selected type 2 cytokines (y-axis, pg/ml) in (B) nasal secretion and (C) sera of allergic (black rhombus, *n*=12) and non-allergic (white rhombus *n* =12) subjects showing no significant differences using Mann-Whitney U test (*p*>0.05). Lines represent the median with a 95% confidence interval and the standard error of the median.

### 2.4. Cytokine measurements

Thirty-three cytokines were analysed in nasal secretions and sera for using the Meso Scale Discovery (MSD) multiplex U-Plex platform (MSD, Rockville, MA, USA) as described before ^20^, and as outlined in the online repository.

### 2.5. Mass Cytometry: Staining and acquisition of PBMCs using MaxPAR® Immunoprofiling Assay and CyToF

Before immunostaining PBMCs, the frozen cells underwent a careful thawing process, as described in the online repository. For immunostaining, the PBMCs were subjected to the MaxPAR Direct Immunoprofiling Assay (Standard BioTools, South San Francisco, CA, USA) supplemented with commercial and in-house labeled antibodies given in Supplementary Table E3 following the manufacturer’s instructions. A brief description of the staining and acquisition procedure can be found in the online repository. A detailed protocol has been described elsewhere ^21^.

### 2.6. Processing of Mass Cytometry Data Using Dimensionality Reduction and FlowSOM

A step-wise approach was followed for analysis: normalized raw data files were imported into Cytobank (Beckman Coulter, Brea, CA, USA), and then quality control was employed to remove undesired events (dead cells, debris, normalization beads, aggregates, and coincident ion clouds) by manually gating out beads and according to residual, center, offset, width, event length, and DNA intercalator signals in biaxial plots vs Time parameter using Gaussian discrimination, leaving viable cell events for downstream analysis.

After data clean-up, dimensionality reduction was performed using opt-SNE in Cytobank on gated live cells using 38 channels (refer to Supplementary Table E3) with the advanced settings left on the software’s default.

The opt-SNE was then visually inspected to identify distinctive clusters. Then, FlowSOM clustering was performed on Cytobank (Beckman Coulter) utilizing the opt-SNE-reduced data as input. All the channels used for opt-SNE were also implemented in the FlowSOM analysis. Subsequently, clusters were further annotated based on characteristic marker expression patterns. Cell population/cluster percentage data across different groups and markers’ median metal intensity (MMI) were exported to GraphPad prism (GraphPad Software, Boston, MA, USA) for a statistical analysis and graph output. Further information on opt-SNE and FlowSOM algorithm settings can be found on Cytobank’s platform (cytobank.org).

### 2.7. Statistical Analysis

All statistical analyses were conducted using GraphPad Prism 9.5.1(GraphPad Software). Cytokine values underwent a log transformation. Hierarchical clustering analysis of levels of cytokines and immune cell populations was performed using a heat-mapper with Spearman Rank Correlation as a distance measurement method ^22^. Furthermore, data on the percentage of cell populations and marker expression were retrieved from Cytobank and subsequently imported into GraphPad Prism for comprehensive analysis.

Given the nature of the data distribution, non-parametric tests were employed for group comparisons. Specifically, the Mann-Whitney U test was utilized to evaluate differences between allergic and non-allergic individuals. To assess distinctions among the CRSsNP, CRSwNP, and N-ERD groups, the Kruskal-Wallis test was applied along with Dunn’s test for multiple comparisons. For all analyses, a significance level of *p* < 0.05 was established as the threshold for statistical significance.

## 3. RESULTS

### 3.1. Subject characteristics

Demographic and clinical characteristics of the study cohorts are given in Table I. A total of 24 participants grouped into subjects without any signs of acute or chronic nasal inflammation=Controls (*n=6*), CRSsNP (*n*=6), CRSwNP (*n*=6) and N-ERD (*n*=6) were included in the study. Half of the cohorts in each group were known to have a respiratory allergy (*n=*3 per group) or were not allergic (*n=*3 per group). An individual overview of the patient characteristics is provided in Supplementary Table E1.

### 3.2. Immunological profiling of allergic and non-allergic cohorts based on allergen-specific IgE and cytokine levels

Participants were categorised as allergic or non-allergic based on their clinical history and allergen-specific positive Skin Prick Test reaction or specific serum IgE levels by ALEX2 microarray. As shown in Fig. 1A, all patients (except two individuals with positive skin prick test only) classified as allergic based on their clinical history showed detectable (≥0.35 kUA/L) specific serum IgE levels to at least one respiratory allergen.

Levels of 33 cytokines measured both in nasal secretions and serum showed no significant differences between allergic and non-allergic subjects (data not shown); exemplary type 2 cytokine levels are given for nasal secretions (Fig. 1B) and serum (Fig. 1C).

### 3.3. Cytokine profiling reveals distinct inflammatory patterns in nasal secretions of N-ERD, CRSsNP, and CRSwNP patients

To determine differences in cytokine patterns in patients suffering from various CRS disease entities compared to disease controls, hierarchical clustering analysis of cytokine levels in nasal secretion (Fig. 2A) and serum (Fig. 2B) was performed. In nasal secretions, we found a clustering of type 2 associated cytokines CCL17, Eotaxin, Eotaxin-3, IL-5 and IL-13, which were highly upregulated in N-ERD patients but also elevated in CRSwNP patients. Individual cytokine analysis confirmed that N-ERD patients showed significantly elevated levels of IL-5 compared to the Control (*p=*0.0423) and CRSsNP (*p=*0.0423) groups. Similarly, there were significantly higher levels of IL-9 (*p*=0.0018), CCL17 (*p*=0.0225), Eotaxin (0.0076), and Eotaxin-3 (*p*=0.0049) in N-ERD patients in comparison to CRSsNP (Fig.2C). In contrast to nasal secretions, no significant differences in cytokine levels were observed in sera of the patients between groups (Fig. 2D).

**Figure 2:**
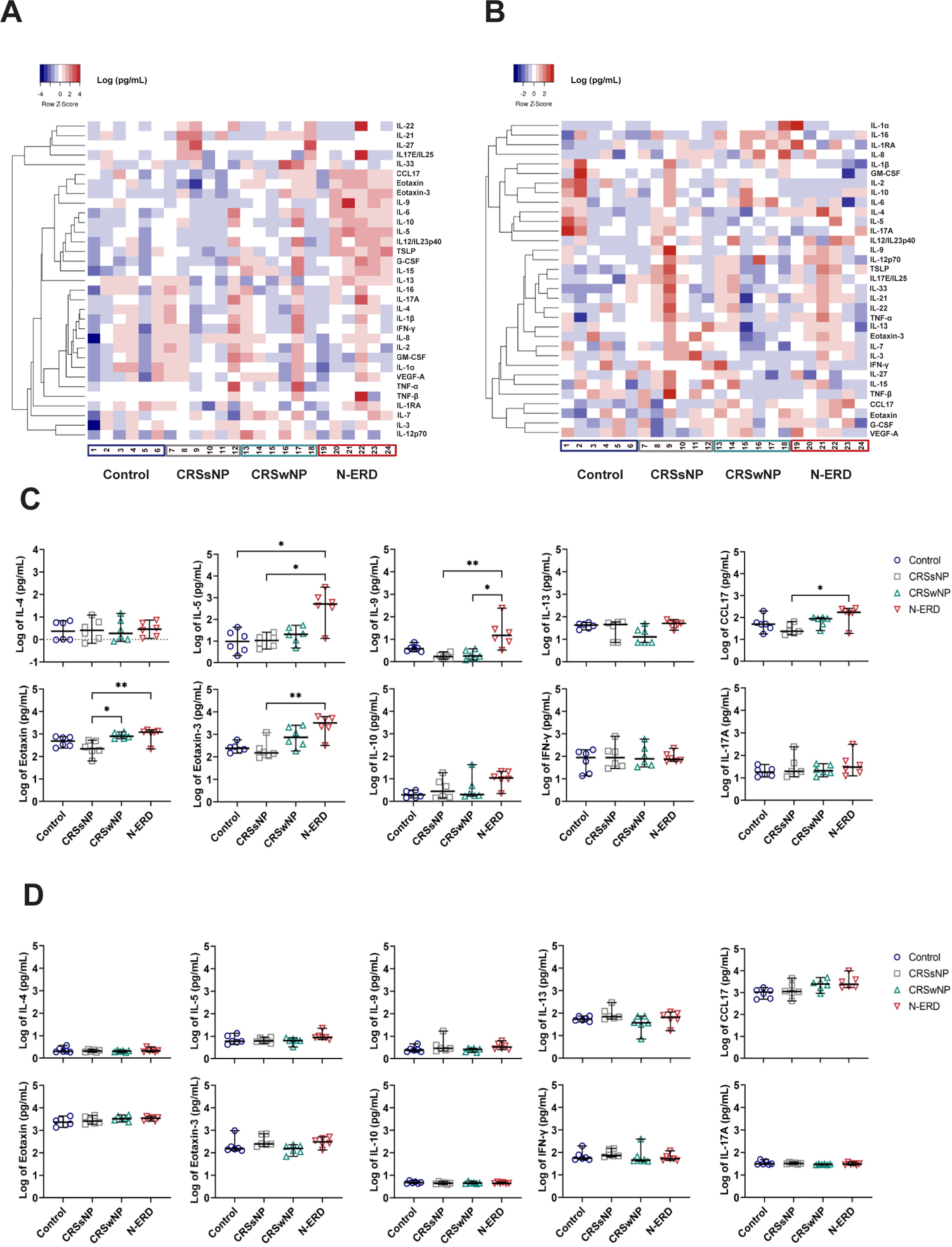
Elevated levels of type 2 cytokines in nasal secretions but not serum of patients suffering from chronic rhinosinusitis (CRS) with nasal polyps (wNP) and from nonsteroidal anti-inflammatory drugs (NSAIDs)-exacerbated respiratory disease (N-ERD). (A, B) Cytokine heat map and dendrogram of hierarchical clustering analysis of expression levels of 33 cytokines in (A) nasal secretion and (B) serum of Controls (*n*=6), CRS without nasal polyps (CRSsNP, *n*=6), CRSwNP (*n*=6) and N-ERD *(n*=6). The color gradient from blue to red represents an increasing quantity of cytokines (pg/ml). (C, D) Levels (y-axis, log of pg/ml) of interleukin (IL)-4, IL-5, IL-9, IL-13, Chemokine (C-C Motif) Ligand (CCL)17, Eotaxin, Eotaxin-3, IL-10, interferon (IFN)-γ and IL-17A in (C) nasal secretions and (D) serum in Controls (blue circle, *n*=6), CRSsNP (grey square, *n*=6), CRSwNP (green triangle, *n*=6) and N-ERD (red triangle, *n*=6) are displayed. Lines represent the median with a 95% confidence interval and the standard error of the median; stars represent statistically significant differences using the Kruskal-Wallis test followed by Dunn’s test (*: *p*≤0.05, **: *p*≤0.01).

### 3.4. High-resolution immune profiling by mass cytometry reveals significantly higher numbers of Th2a cells in the blood of allergic subjects but not of patients suffering from CRS

To get an insight into the immune cell subsets associated with CRS as well as allergic diseases, high-resolution immune profiling was conducted in PBMCs of the participants (*n=22*) by mass cytometry. Two patients in the CRSwNP allergic group were excluded from the analysis due to insufficient cell numbers for in-depth analysis. Mass cytometry data, coupled with dimensionality reduction techniques (Fig. 3A) and unbiased automated clustering analyses by FlowSOM, was conducted and showed clear separation of cell populations based on canonical markers as exemplified in Fig. 3A. As shown in Fig. 3B and Fig. 3C clusters of immune cells, encompassing natural killer (NK), natural killer T (NKT), mucosal-associated invariant T (MAIT), CD4+ T (CD4 T), Regulatory T (T-reg), CD8+ T (CD8 T), Gamma-Delta T (γδ T), Memory B (Memory B), Naive B (Naive B), Plasmablasts (Plasmas), Monocytes, myeloid Dendritic cells (mDCs), and plasmacytoid Dendritic cells (pDCs) in the different disease entities. Differential abundance analysis to quantify immune cell subpopulations revealed only a slight trend towards increased levels of T-reg and Th2 cells in the PBMCs of CRSsNP and N-ERD patients (Supplementary Fig. E1). No significant alterations in the other cell types were identified between the disease groups (Fig. 3D).

**Figure 3:**
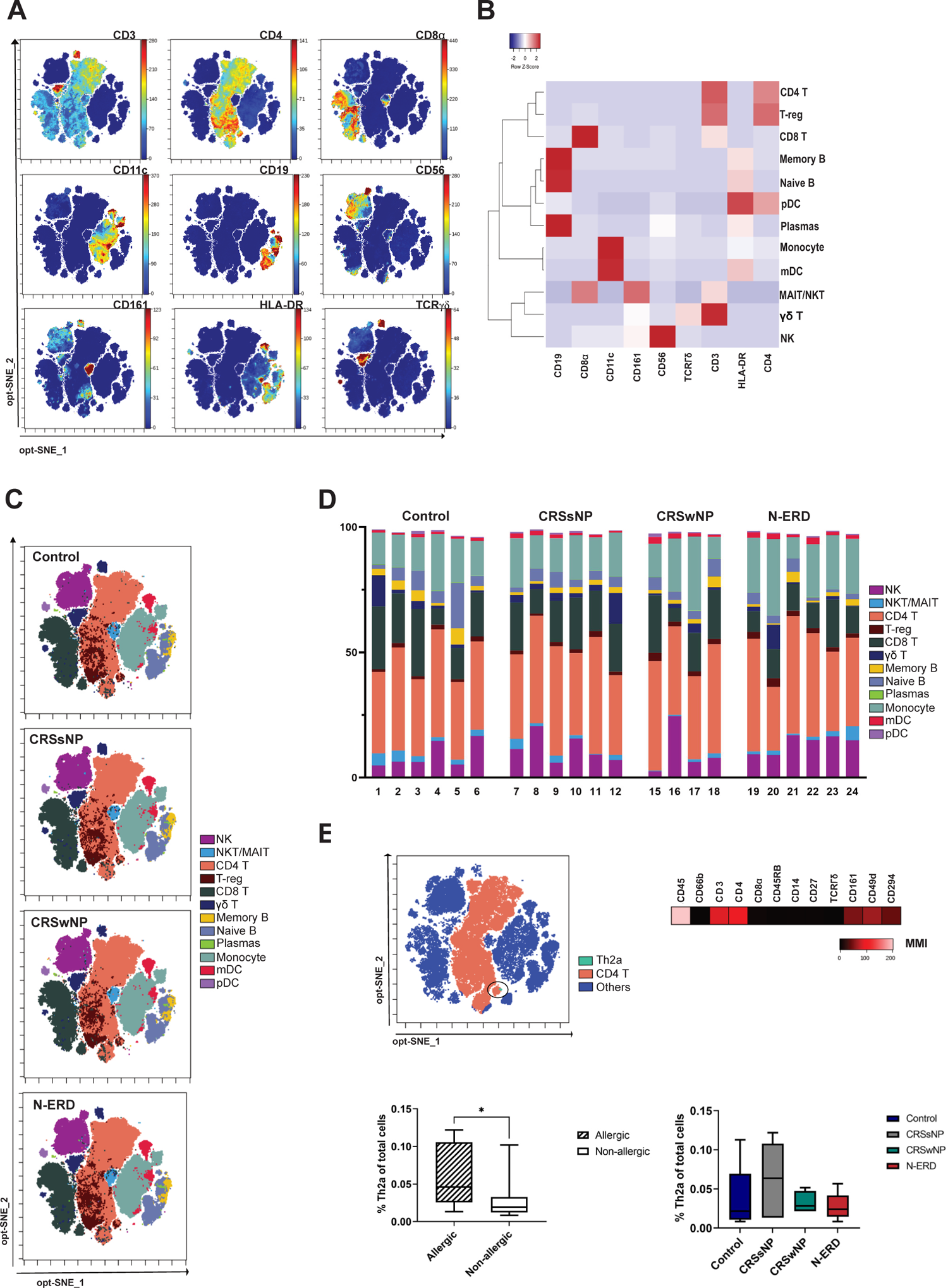
Mass cytometry analysis of peripheral blood mononuclear cells (PBMCs) in control, chronic rhinosinusitis (CRS) without nasal polyps (CRSsNP), CRS with nasal polyps (CRSwNP) and Nonsteroidal anti-inflammatory drugs (NSAIDs)-Exacerbated Respiratory Disease (N-ERD) patients. (A) An optimized t-distributed stochastic neighbour embedding (opt-SNE) map representing a consensus staining pattern and intensity levels of key markers employed in mass cytometry, specifically, the markers CD3, CD4, CD8, CD11c, CD19, CD56, CD161, HLA-DR, and TCRγδ depicted in a concatenated file. The color gradient from blue to red represents an increasing intensity of the metals tagged to the specific antibodies. (B) Heat map and dendrogram of hierarchical clustering analysis displaying the expression levels/metal intensities of selected markers across individual immune cell types. (C) A consensus overlay of FlowSOM metaclusters on an opt-SNE map of concatenated files and (D) bar chart graph of individual patients displaying major cell populations characterized after FlowSOM analysis. Major immune cell types namely Natural Killer (NK) Cells, Natural Killer T (NKT), Mucosal-associated invariant T (MAIT) Cells, CD4+ T Cells (CD4 T), regulatory T cells (T-reg), CD8+ T Cells (CD8 T), Gamma-Delta T Cells (γ δ T), Memory B Cells (Memory B), Naive B cells (Naive B), Plasmablasts (Plasmas), Monocytes, Myeloid Dendritic Cells (mDCs), Plasmacytoid Dendritic Cells (pDCs) were identified in all four patient groups: controls (*n*=6), chronic rhinosinusitis with (CRSwNP, *n*=6) and without (CRSsNP, *n*=4) nasal polyps and non-steroidal anti-inflammatory drugs (NSAIDs)-exacerbated respiratory disease (N-ERD, *n*=6). (E) Th2a subset of the CD4+ T cells, manually gated (upper left panel) using marker expression patterns shown on a heat map (upper right panel). Percentage Th2a cells (y-axis) of total PBMCs in allergic (lower left panel, *n*=10) versus non-allergic (lower left panel, *n*=12) and in disease groups (lower right panel). Box plots show the range with medians as horizontal lines and whiskers indicating minimum and maximum values. Stars represent statistically significant differences between allergic and non-allergic groups using Mann-Whitney U test (*: *p*≤0.05) whereas the Kruskal-Wallis test followed by Dunn’s test in disease groups showed no significant difference (*p*>0.05).

As Th2a cells have previously been described to be elevated in the blood of allergic subjects^23^, we next investigated if they were also elevated in CRS subtypes with strong type 2 profile. To that aim, we manually gated the subpopulation of Th2a cells based on established surface marker expression (CD27^-^CD45RB^-^CD161^+^ CRTH2/CD294^+^CD49d^+^^24^) within PBMCs (Fig. 3E). Despite being a relatively small fraction of the total PBMC population, the Th2a immune cell subset exhibited a notable increase in allergic individuals (*p*=0.0133).

Intriguingly, when comparing these cells within the CRS groups, no significant changes were observed (*p*=0.7650) (Fig. 3E). As shown in Supplementary Fig. E2, no differences were seen when CRS and allergy status stratified the data.

### 3.5. Comprehensive profiling of B cell subpopulations using FlowSOM analysis

Given the pivotal role of B cells in the pathophysiology of both CRS and allergic conditions, we conducted an extensive analysis of B cell populations (Fig. 4) within PBMCs. This analysis was performed using an automated assessment of marker expression profiles via FlowSOM leading to identification of ten distinct B-cell metaclusters (B1-B10) (Fig. 4A).

**Figure 4:**
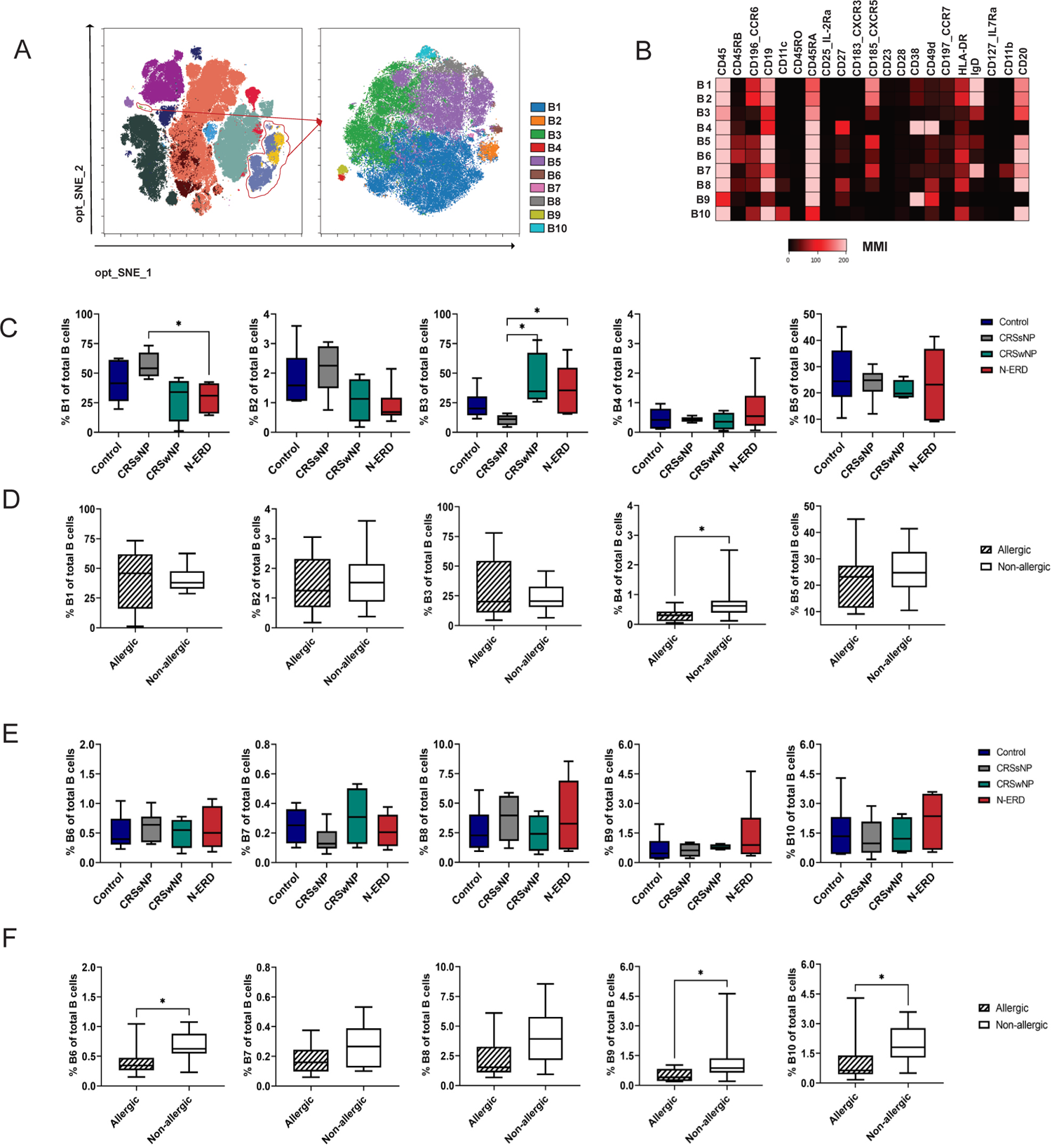
In-detail B cell subset identification using FlowSOM. (A) B cell FlowSOM clusters (left panel, outlined in red lines) were merged into one population and subjected to further sub-clustering using FlowSOM analysis (right panel) into ten subtypes of B cells, labelled B1 to B10. (B) A consensus heatmap of the raw values of median metal intensity (MMI) of selected markers in B1 to B10, the colour gradient from black to red represents an increasing expression/intensity of the markers in the concatenated file. (C-F) Comparison of percentage (y-axis) of respective B cell sub-clusters (B1-B10) of total B cells in (C, E) Disease conditions (controls, blue, *n*=6; CRS without polyps (CRSsNP), grey, *n*=6; CRS with nasal polyps (CRSwNP), green, *n*=6) and Non-steroidal anti-inflammatory drug (NSAID)-exacerbated respiratory disease (N-ERD, *n*=6, red)) and (D, F) allergic conditions (allergic, black-white pattern, *n*=10; non-allergic, white, *n*=12). Box plots show the range with medians as horizontal lines and whiskers indicating minimum and maximum values. Stars represent statistically significant differences between groups using Kruskal-Wallis test (C, E) followed by Dunn’s test (*: *p*≤0.05) and (D, F) using the Mann Whitney U test (*: *p*≤0.05).

Notably, several markers, including CD45RB, CCR6, CD11c, CD27, CXCR5, CD38, CD49d, HLA-DR, CD11b, and CD20 exhibited differential expression within the B cell population, giving rise to these uniquely defined metaclusters (Fig. 4B).

Broadly, the metaclusters consisted of naive B cells, characterized by CD27^-^ (Fig. 4B) (specifically B1, B2, B3, B7 & B10), and memory B cells, characterized by CD27^+^ (including B4, B5, B6, B8 & B9), among which B4 and B9 displayed high CD38^+^ expression and thus are putative plasmablasts. As illustrated in Fig. 4C-F, the metaclusters were differently sized, with B1, a naïve B cell metacluster, constituting approximately 60% of the total B cell population in control subjects. In contrast, B7, another type of naïve B cell distinguished by CD11b surface marker, accounted for just 0.4% of the B cell population in control subjects.

To investigate potential differences, we conducted an abundance analysis of the identified B cell metaclusters across various groups: Control, CRSsNP, CRSwNP, and N-ERD, as well as allergic and non-allergic subjects (Fig. 4C-F). Notably, metaclusters B1 and B2 (representing naïve B cells marked by higher expression of CCR6 and CXCR5) showed a trend towards increased levels in CRSsNP patients, which reached statistical significance for B1 cells as compared to N-ERD patients (*p*=0.0231). Conversely, N-ERD and CRSwNP groups showed a significantly increased frequency of B3 (naïve B cell with low expression of CCR6) compared to CRSsNP (N-ERD-CRSsNP: *p*=0.0141, CRSwNP-CRSsNP: *p=*0.0266) (Fig. 4C).

A more in-depth analysis of the abundance in the context of allergy (Fig. 4D-F; Supplementary Fig. E3) revealed that B3 was not increased in allergic subjects, indicating that the observed increase in the naïve B1, B2, and B3 frequency was solely due to the underlying CRS disease. Allergic subjects, however, had a decreased frequency of memory B-cell metaclusters B4 (*p*=0.0169), B6 (*p*=0.0206), B9 (*p=*0.0426), and naïve B cell metacluster B10 (*p*=0.0249), as compared to non-allergic subjects.

### 3.6. Immunophenotypic distinctions in B cell subtypes illuminate unique features of N-ERD derived B cells

We further analysed surface marker expression levels in B cell metaclusters of the three CRS disease groups. N-ERD and CRSsNP patients showed a trend towards elevated median metal intensity (MMI) levels of the differentiation marker CD45RA across all B cell metaclusters, which reached a significant difference in B10 of CRSsNP compared to Control (*p=*0.0082).

Conversely, N-ERD patients displayed decreased levels of CXCR5 expression in all B cell metaclusters B1 to B10 when compared to Control, CRSsNP, and CRSwNP groups (Fig. 5B), yet differences were not statistically significant (*p>*0.05). Similarly, N-ERD patients exhibited a significantly decreased expression of CD49d (*p=* 0.0402) in B9 metacluster relative to the Control (Fig. 5B). Overall, these findings highlight distinct immunophenotypic changes in B cell subtypes in N-ERD patients, characterized by elevated CD45RA expression and reduced levels of CXCR5 and CD49d.

**Figure 5:**
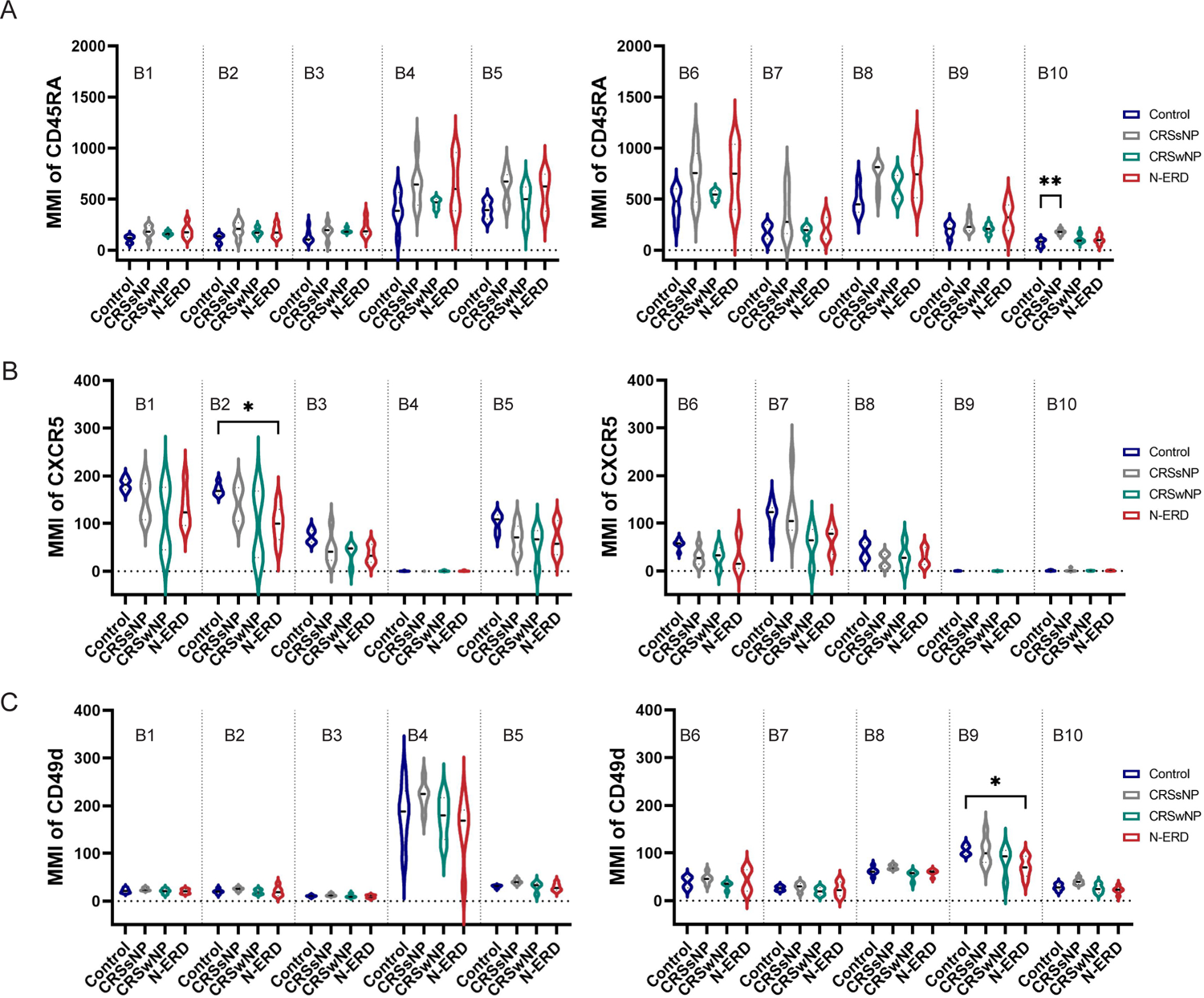
CD45RA, CXCR5 and CD49d expression levels in B cell subtypes. (A-C) Median metal intensity (MMI, y-axis) in B cell sub-clusters B1-B5 (left panels) and B6 to B10 (right panels) of (A) CD45RA, (B) CXCR5 and (C) CD49d in controls (blue, *n*=6), chronic rhinosinusitis without (CRSsNP, grey, *n*=6) and with (CRSwNP, green, *n*=4) nasal polyps and in patients with non-steroidal anti-inflammatory drugs (NSAIDs)-exacerbated respiratory disease (N-ERD, red, *n*=6). Stars represent statistically significant differences between groups using Kruskal-Wallis test followed by Dunn’s test (*: *p*≤0.05).

Furthermore, we analysed the MMI in B cell metaclusters in allergic and non-allergic individuals. The expression levels were comparable between the two groups across most B cell metaclusters, except for B7, where expression levels of CD49d were significantly higher in non-allergic as compared to allergic patients (*p*= 0.0138) (Supplementary Fig. E4).

### 3.7. Heterogeneity in Natural Killer (NK) Cell Populations in CRSsNP, N-ERD, and allergic individuals

Another focus of this study was exploring the heterogeneity of Natural Killer (NK) cell populations in the study cohorts as NK cell dysfunction has previously been reported in CRS^25^. The NK metaclusters were characterized based on their surface marker expression profiles (Fig. 6A and B). They revealed four distinct NK metaclusters, Late-stage (LS) NK1, LS NK2, Early-stage (ES) NK1, and ES NK2, in the PBMCs. While no significant differences in abundance were observed between the CRSsNP, CRSwNP, N-ERD, and Control (*p*>0.05) (Fig. 6C), an increased frequency of LS NK2 cells was noted in non-allergic individuals compared to allergic individuals (*p*=0.0215). Additionally, expression levels of CD45RA and CD56 within different NK metaclusters exhibited variations in the context of CRSsNP, CRSwNP, and N-ERD (Fig. 6E). Overall, both CRSsNP and N-ERD groups demonstrated elevated levels of CD45RA across all metaclusters, with LS NK1 exhibiting a statistically significant increase in CRSsNP versus control (*p*=0.0306). Similarly, CRSsNP groups showed an elevated level of median CD56 intensity, which showed significant differences in ES NK 1 cells compared to CRSwNP patients (*p*=0.0323).

**Figure 6:**
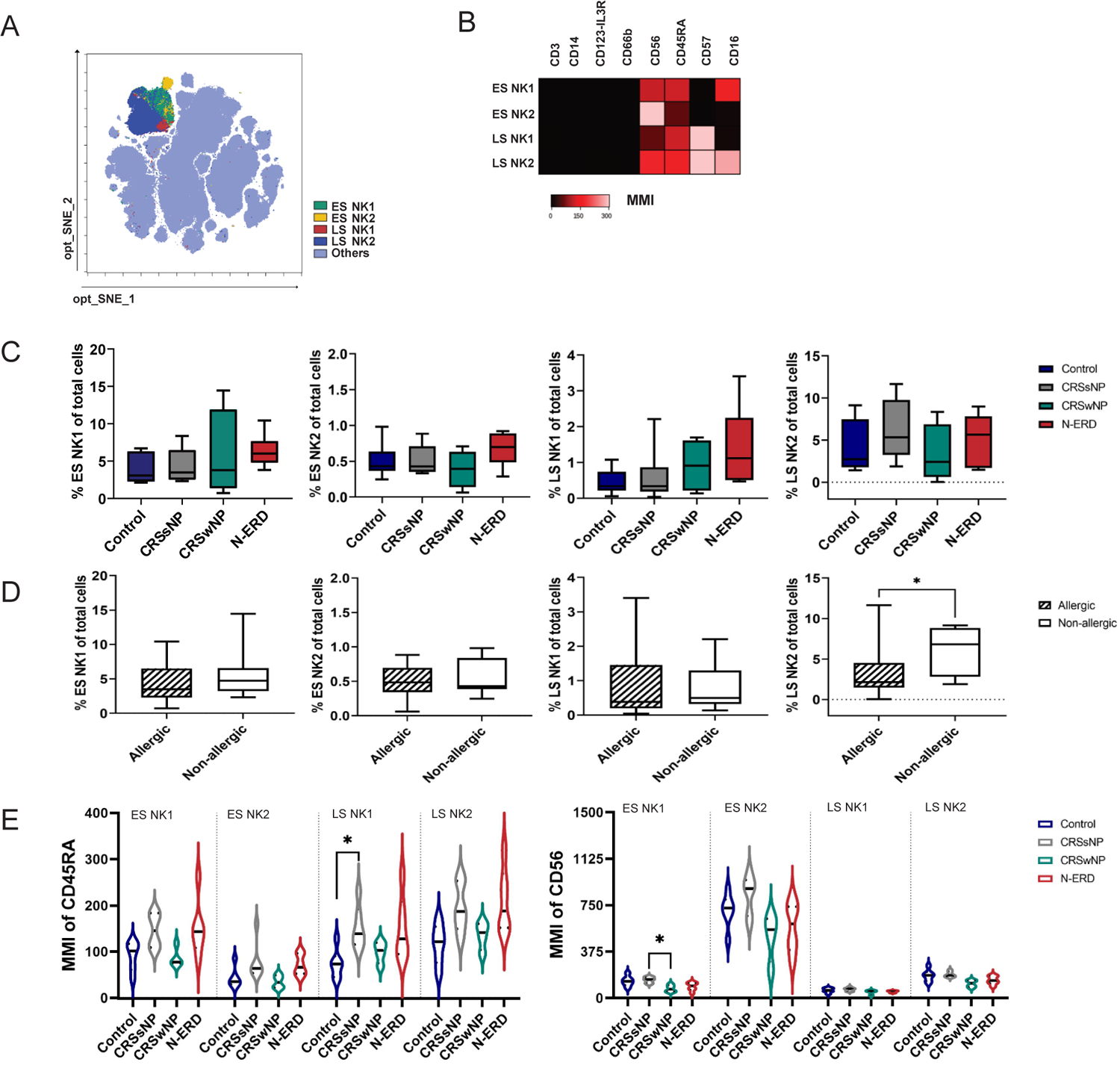
Characterization of early-stage (ES) and late-stage (LS) Natural killer cells (NKs). (A) A consensus optimized t-distributed stochastic neighbour embedding (opt-SNE) map showing ES and LS NK cells. (B) A consensus heatmap of the raw median metal intensity (MMI) values of selected markers characterizing NK cell subsets. The color gradient from black to bed represents an increasing expression/intensity of the markers. (C, D) Percentage of NK cell subsets of total cells (y-axis) in (C) disease groups (Controls, blue, *n*=6; chronic rhinosinusitis (CRS) without nasal polyps (CRSsNP), grey, n=6; CRS with nasal polyps (CRSwNP), green, *n*=4; non-steroidal anti-inflammatory drugs (NSAIDs)-exacerbated respiratory disease (N-ERD), red, *n*=6) and in (D) allergic (black-white pattern, *n*=10) and non-allergic (white, *n*=12) patients. (E, F) Median metal intensity (MMI) of (E) CD45RA and (F) CD56 in NK cell subtypes in disease groups (as described in C). Box plots show the range with medians as horizontal lines and whiskers indicating minimum and maximum values. Stars represent statistically significant differences between groups using Kruskal-Wallis test (C, E) followed by Dunn’s test (*: *p*≤0.05) and (D) using Mann-Whitney U test (*: *p*≤0.05).

## 4. DISCUSSION

Here, we conducted a detailed analysis of immune cell profiles in PBMCs of patients suffering from various forms of CRS: CRSsNP, CRSwNP, and N-ERD, with a further distinction in patients with and without respiratory allergies.

Using a panel comprising 33 different cytokines, we found elevated concentrations of type 2-associated cytokines, including CCL17, Eotaxin, Eotaxin-3, IL-5, and IL-9, in nasal secretions of CRSwNP patients with the highest levels observed in N-ERD patients. This agrees with earlier reports of CRSwNP and especially N-ERD being dominated by type 2-driven inflammatory conditions in the Western world ^2,4–6^. CRSsNP patients showed a more mixed pattern of type 1 and type 3 endotypes and strong inter-individual differences in accordance with previous reports ^2,4–6^.

Our main objective in this study was to investigate differences in levels and characteristics of various immune cell types within CRS using mass cytometry. With regards to T cell subpopulations, we detected no significant differences in peripheral blood. Though mass cytometry has recently revealed differences in nasal polyp-derived T cell subsets with a higher percentage of CD161^hi^ CXCR3^lo^ expressing T helper cells in nasal polyps as compared to control tissues ^26^, we only detected a trend towards elevated Th2 cell levels in blood of CRSwNP patients and N-ERD patients, which did not reach significance. The lack of difference in Th2 cell numbers between different patient groups has also been observed using flow cytometry approaches ^27^.

To the best of our knowledge, we are the first to identify Th2a cells based on the expression of CRTH2, CD161, CD49d and CD27 in the blood of patients using mass cytometry. This Th2 subset was initially described by Wambre et al as associated with allergic disease status ^23^ and recently put forward as a candidate for monitoring success during immunotherapy for food ^28^ or respiratory allergens ^29^. Interestingly, we found no differences in Th2a levels between CRS entities but significantly increased levels in allergic versus non-allergic patients regardless of their CRS status. Our observation that allergic patients had significantly elevated levels of Th2a cells despite being a heterogeneous group with various levels of specific IgE against different allergen sources, from tree to animal, implicates Th2a levels as a potentially universal biomarker and therapeutic target for allergic disease ^30^.

Eosinophils have long been regarded as main drivers of CRSwNP. However, with the introduction and success of anti-IgE and anti-IL4/IL-13 antibodies in treating severe disease^31^, B cells have also been attributed to an important role in pathogenesis ^32, 33^. In this line, it has been shown that chronic airway inflammation creates an environment well-suited for B cell differentiation ^34^ and that the nasal mucosa of CRS shows elevated levels of naïve B cells, memory B cells and plasma cells, with the highest number observed in CRSwNP patients ^35,36^. In the blood of CRS patients, we did not observe differences in major B cell subsets, which is in agreement with previous data ^35^. Thus, we performed a deeper characterization employing an unsupervised high-resolution profiling approach using FlowSOM. This enabled us to distinguish ten distinct B cells subtypes based on the expression of phenotypic markers, including CD45RB, CD27, CXCR5, CD38, CD49d, HLA-DR, CD19, and CD20. Though not significant, we observed in CRSsNP patients a trend towards elevated levels of B1 and B2 metaclusters, which show characteristics of resting naïve B cells (CD27^-^CD38^+^IgD^+^) ^37^. N-ERD patients, on the other hand, displayed a significantly increased frequency of B3, another subset of naive B cells characterized by the absence of CD27 and CD38 expression but positive for IgD. These cells can be identified as putative “mature-naive” ^38^ or “activated naïve” ^37^ B cells. Additionally, B-cells of CRS patients showed a trend towards reduced levels of the chemokine receptor CXCR5 and the integrin CD49d in selected metaclusters, which are both involved in establishing and maintaining optimal germinal center reactions ^39,40^. These findings are interesting in light of previous observations that IgE-secreting cells from nasal polyps are mainly guided to extrafollicular pathways ^34,41^. In summary, it is compelling to speculate that blood-borne activated naïve B cells, enriched in N-ERD patients, may migrate to extrafollicular structures within the nasal polyp, where they may further differentiate into antibody-producing cells ^33^.

NK cell dysfunction has also been observed in CRS patients with decreased effector function being associated with increased eosinophilic inflammation ^42^ and disease severeness ^43^. In this context, we observed reduced expression of the phenotypic NK cell activation marker CD56 ^44,45^ in CRSwNP patients. As CD56-negative NK cell subsets are enriched in patients with chronic virus infections ^46,47^, it is conceivable that the loss of CD56 observed in our study could be at least partly driven by the repeated infections in the course of CRS.

In patients suffering from respiratory allergies, we observed significantly reduced levels of late-stage NK cells expressing CD16 regardless of their allergic status. However, there were no differences in the CD56^+^CD16^-^ subsets as previously described in peanut-allergic subjects ^48^ or subjects suffering from atopic dermatitis ^49^. Our differential results could be both due to the different routes of allergen sensitization or due to the absence of allergen restimulation in our experimental design as compared to Zhou et al.^48^

Limitations of our study include the relatively small sample size with regard to different CRS subtypes and the absence of nasal mucosa samples. It could also be that the often discussed lower sensitivity of mass cytometry ^50^ as compared to flow cytometry may have had an impact on our results. However, a recent report showed the comparability between the two techniques with regard to T cell subsets ^26^.

In conclusion, this study provided a comprehensive analysis of blood-derived immune cell profiles in various forms of CRS and allergic patients. Notably, an increased presence of Th2a cells, pivotal in allergic responses, was observed in allergic cohorts. Further, our analysis of B and NK cell populations identified distinct subtypes with potential implications for both type 2 diseases. These findings contribute to a deeper understanding of the immunological profiles in PBMCs of patients with CRS or allergy, shedding light on potential avenues for further research and therapeutic approaches.

## ABBREVIATIONS

CCL: Chemokine (C-C Motif) Ligand

CD: Cluster of differentiation

CRS: Chronic rhinosinusitis

CRSsNP: Chronic rhinosinusitis without nasal polyps

CRSwNP: Chronic rhinosinusitis with nasal polyps

CyTOF: Cytometry by Time-Of-Flight

G-CSF: granulocyte-colony stimulating factor

GM-CSF: Granulocyte-macrophage colony-stimulating factor

Ig: Immunoglobulin

IL: Interleukin

IFN: Interferon

MMI: Median metal intensity

NSAIDs: Non-steroidal anti-inflammatory drugs

N-ERD: NSAIDs-exacerbated respiratory disease

opt-SNE: Optimized t-distributed stochastic neighbour embedding

PBMC: Peripheral blood mononuclear cells

SNOT-20: 20-item sinonasal outcome test

TNF: Tumour necrosis factor

TSLP: Thymic stromal lymphopoietin

VEGF: Vascular endothelial growth factor

## Data Availability

All data produced in the present study are available upon reasonable request to the authors

## ACKNOWLEDGMENT

We would like to thank the technical teams of Standard BioTools and Cytobank for their support during the study.

