## Supplementary Material for "Deep immune profiling of chronic rhinosinusitis in allergic and non-allergic cohorts using mass cytometry"

1. **Supplementary Methods**
   1.1. *Sera*

Blood was collected in a VACUETTE® Clot Activator tube (Greiner Bio-One International GmbH, Frickenhausen, Germany) and left undisturbed at room temperature for 30 minutes to clot. Subsequently, the tube was centrifuged at 1790 × g for 10 minutes at room temperature. The resulting sera were aliquoted into 2 mL screw top vials and stored at -80°C until used for subsequent analyses.

1.2. *Nasal secretion*

Nasal secretions samples were obtained from both nostrils of all participants using the Nasosorption™ FX·I (Hunt Developments, Midhurst. UK) nasal sampling device. Nasosorption was performed by gently inserting two strips of synthetic absorptive matrix into the nostrils of the participants and pressing against the inferior turbinate for 2 minutes. Subsequently, the strips were carefully removed and placed into a Spin-X filter Eppendorf tube containing 300 μl of elution buffer with PBS/1% Bovine Serum Albumin (Thermo Fisher Scientific, Waltham, Massachusetts, USA). The mixture was then vortexed for 30 seconds. The strips were transferred to a filter cup within a microcentrifuge tube and centrifuged for 20 minutes at 16,000 × g at 4°C. The supernatant aliquots were then stored at −80°C until further use.

1.3. *PBMCs*

Fresh whole blood (9 mL) was collected by standardized venipuncture Heparinized VACUETTE® tubes (Greiner Bio-One International GmbH) from which PBMCs were then isolated by centrifugation on a density gradient of Ficoll (Ficoll® Paque Plus, MilliporeSigma, Darmstadt, Germany). The PBMCs were resuspended in PBS for washing. Then pellets were resuspended in a cryoprotective solution composed of Fetal Bovine Serum (FBS) (Thermo Fisher Scientific, Massachusetts, USA) and 10% dimethyl sulfoxide (DMSO) (MilliporeSigma) at 1-2x10^7^ cells per mL into a Cryovial, under sterile conditions. The cells were then transferred to a controlled-rate freezer, gradually lowering the temperature to Liquid Nitrogen (-180°C) until further use.

1.4. *IgE measurement*

The measurement was performed using the ALEX® Allergy Explorer assay (Allergy Explorer version 2, MacroArray Diagnostics, Vienna, Austria), according to the manufacturer's instructions. Briefly, 400µL sample diluent and 100µL thawed patient serum were added to the array and incubated for 2 hours at room temperature with agitation. Subsequently, 500µL of detection antibody was added, incubated for 30 minutes, and returned to the shaker at room temperature. After this, 500µL of substrate was introduced and incubated for 8 minutes. Finally, the reaction was stopped by adding 100µL of Stop Solution. Both total and allergen-specific IgE levels were quantified using the RAPTOR software (MacroArray Diagnostics).

1.5. *Cytokine measurements with MSD*

The measured cytokines include CCL17, Eotaxin, Eotaxin 3, G-CSF, GM-CSF, IFN-γ, IL-10, IL-12/IL23p40, IL-12p70, IL-13, IL-15, IL-16, IL-17A, IL-1α, IL-1RA, IL-1β, IL-2, IL-3, IL-4, IL-5, IL-6, IL-7, IL-8, IL-9, IL-21, IL-22, IL17E/IL-25, IL-27, IL-33, TNF-α, TNF-β, TSLP, and VEGF-A. Nasal secretions and serum samples were thawed, and nasal secretions from both nostrils were pooled before application to MSD plates. Measurements were conducted according to the manufacturer's instructions(<https://www.mesoscale.com/en>). Briefly, linkers were complexed with target antibody, diluted, and coated onto plates. Standards were freshly reconstituted and serially diluted. Samples and standards were incubated on the plate overnight at 4°C. Detection antibodies were applied, followed by washing and immediate measurement. Data acquisition was performed using DISCOVERY WORKBENCH software (MSD), with both standards and samples measured in duplicate. Cytokine values below the level of assay detection were replaced by the values representing the lower detection limit.

1.6. *Mass Cytometry*: *Staining of PBMCs using MaxPAR® Immunoprofiling Assay*

Before immunostaining of PBMCs, the frozen cells were thawed as follows. A thawing medium was prepared using Gibco’s RPMI 1640 (Thermo Fisher Scientific, Massachusetts, USA) supplemented with 10% Fetal Bovine Serum (Thermo Fisher Scientific) and pre-warmed to 37°C. A cryovial containing the frozen PBMCs was swiftly thawed in a 37°C water bath and gently transferred from the cryovial into a conical tube containing the pre-warmed thawing medium. The cell suspension was then centrifuged at 300xg for 5 minutes, and the supernatant was aspirated, leaving the pellet undisturbed.

For immunostaining, PBMCs were subjected to the MaxPAR Direct Immunoprofiling Assay (Standard BioTools, South San Francisco, USA) supplemented with commercial and in-house labelled antibodies as detailed in Table E2 following the manufacturer's instructions. Briefly, PBMCs were resuspended in 1 mL of cell staining buffer (CSB) (Standard BioTools) containing 0.5 µl of Nuclease Pierce (MilliporeSigma, Massachusetts, USA) and then incubated for 30 minutes at 37°C in a shaking incubator. Following digestion with Nuclease (MilliporeSigma), cells were washed with 1 mL of CSB by centrifugation at 300× g for 7 minutes. The cells were then blocked in a Heparin solution (MilliporeSigma). Fc receptors were blocked by adding 5 μl of Human TruStain FcX™ to 3 × 10^6^ cells in 50 μl of CSB and incubated for 10 min. About 215 μl of CSB was added to each dried MAXPAR antibody tube and transferred to the PBMCs for antibody staining. After a 30-minute incubation, cells were washed twice in CSB at 300× g for seven minutes. The cells were then fixed in 1.6% paraformaldehyde (Thermo Fisher Scientific) at room temperature for 10 minutes. After removing the paraformaldehyde with centrifugation at 800× g for 7 minutes, the pellet was resuspended in an intercalating isotope, Cell-ID™ Intercalator-Ir, in 1 mL of FIX-PERM solution (Standard BioTools) and incubated for 30 minutes. Until further processing for acquisition, cells were stored at -80°C for up to one month.

1.7. *Acquisition and processing of mass cytometry data*

Samples were acquired on a Helios® mass cytometer using a wide bore injector (Standard BioTools). Before starting the acquisitions, the instrument was prepared and tuned according to manufacturer‘s recommendations. Sample preparation and acquisition was performed as previously described ^1, 2^. Briefly, cells were washed with CSB followed by Cell Acquisition Solution (CAS) (all Standard BioTools), with a final concentration of 5x10^5^ – 1x10^6^ cells/mL. After determining the cell numbers, samples were resuspended in CAS containing 10% of EQ Four Element Calibration Beads (Standard BioTools) and filtered through a 30μm cell strainer snap cap (Corning Incorporated, Corning, New York, United States). Samples were run with an event rate of 100- 300 events/s. The raw data was collected in flow cytometry standard (FCS) files. Signal intensities were normalized using the FCS processing tool integrated in the CyTOF software v7.0 (Standard BioTools) to account for instrument performance and signal drift changes.

2. **Supplementary Tables and Figures**

Supplementary Table E1: Individual Patient Characteristics

| Code | CRS Group | Respiratory Allergy | TPS* | SNOT-22** | Allergy Symptoms^+^ | Asthma | Total Serum IgE | MC^++^ |
| --- | --- | --- | --- | --- | --- | --- | --- | --- |
| 1 | Control | Yes | 0 | 14 | C, R | No | 54 | Yes |
| 2 | Control | Yes | 0 | n.k. | n.k. | n.k | 207.9 | Yes |
| 3 | Control | Yes | 0 | 5 | R | No | 176.9 | Yes |
| 4 | Control | No | 0 | 1 | none | No | < 20 | Yes |
| 5 | Control | No | 0 | 0 | none | n.k | < 21 | Yes |
| 6 | Control | No | 0 | 0 | none | No | < 22 | Yes |
| 7 | CRSsNP | Yes | 0 | 13 | R | No | 767 | Yes |
| 8 | CRSsNP | Yes | 0 | 15 | R | Yes | 57 | Yes |
| 9 | CRSsNP | Yes | 0 | 13 | C, R | No | < 20 | Yes |
| 10 | CRSsNP | No | 0 | 29 | none | n.k | < 20 | Yes |
| 11 | CRSsNP | No | 0 | n.k. | none | No | < 20 | Yes |
| 12 | CRSsNP | No | 0 | n.k. | none | No | < 20 | Yes |
| 13 | CRSwNP | Yes | 4 | 15 | C, R | No | 37.8 | No |
| 14 | CRSwNP | Yes | 4 | 34 | A, C, R | No | 65.3 | No |
| 15 | CRSwNP | Yes | 2 | 30 | A, C, R | No | 35.4 | Yes |
| 16 | CRSwNP | No | 8 | 47 | none | No | 48 | Yes |
| 17 | CRSwNP | No | 3 | 30 | none | Yes | 67.6 | Yes |
| 18 | CRSwNP | No | 6 | 37 | none | No | 45.8 | Yes |
| 19 | N-ERD | Yes | 3 | 17 | A, C, R | Yes | 1561 | Yes |
| 20 | N-ERD | Yes | 8 | 17 | A, C, R | Yes | 244 | Yes |
| 21 | N-ERD | Yes | 7 | 46 | A, C, R | Yes | 178 | Yes |
| 22 | N-ERD | No | 3 | 18 | none | Yes | 363 | Yes |
| 23 | N-ERD | No | 5 | 20 | none | Yes | 81.9 | Yes |
| 24 | N-ERD | No | 6 | 52 | none | Yes | 47.3 | Yes |

*TPS: Total Polyp Score

** Sino nasal Operation Outcome Test

+C, Conjunctivitis; R, Rhinitis; A, Asthma; ++MC, Mass cytometry

n.k. not known

Supplementary Table E2: List of selected allergens examined for allergen-specific serum IgE level measurement of individual participants

| **Allergen** | **Name/Source** |
| --- | --- |
| Aln g 1 | Alder |
| Alt a 1 | Alternaria alternata |
| Art v 1 | Mugwort |
| Bet v 1 | Silver birch |
| Can f 3 | Dogs |
| Cry j 1 | Sugi |
| Cup a 1 | Cypress |
| Der p 1 | Dust mite |
| Der p 2 | Dust mite |
| Der p 23 | Dust mite |
| Fel d 1 | Cats |
| Fel d 2 | Cats |
| Fel d 4 | Cats |
| Fra e 1 | Ash |
| Ole e 1 | Olive |
| Phl p 1 | Timothy grass |
| Phl p 2 | Timothy grass |
| Phl p 5 | Timothy grass |
| Phl p 6 | Timothy grass |

Supplementary Table E3: Mass cytometry panel of antibodies

| Metal conjugate | Antibody/marker | Clone | Manufacturer |
| --- | --- | --- | --- |
| 89Y | CD45 | HI30 | Standard Biotools |
| 110Cd | CD86 | BU63 | Biolegend (custom conjugated |
| 112Cd | CX3CR1 | 2A9-1 | Biolegend (custom conjugated |
| 114Cd | CD45RB | MEM-55 | Biolegend (custom conjugated) |
| 116Cd | FcεRIα | AER-37 (CRA-1) | Biolegend (custom conjugated) |
| 141Pr | CD196 (CCR6) | G034E3 | Standard Biotools |
| 142Nd | CD1c | L161 | Biolegend (custom conjugated) |
| 143Nd | CD123 (IL-3R) | 6H6 | Standard Biotools |
| 144Nd | CD19 | HIB19 | Standard Biotools |
| 145Nd | CD4 | RPA-T4 | Standard Biotools |
| 146Nd | CD8a | RPA-T8 | Standard Biotools |
| 147Sm | CD11c | Bu15 | Standard Biotools |
| 148Nd | CD16 | 3G8 | Standard Biotools |
| 149Sm | CD45RO | UCHL1 | Standard Biotools |
| 150Nd | CD45RA | HI100 | Standard Biotools |
| 151Eu | CD161 | HP-3G10 | Standard Biotools |
| 152Sm | CD194 (CCR4) | L291H4 | Standard Biotools |
| 153Eu | CD25 | BC96 | Standard Biotools |
| 154Sm | CD27 | O323 | Standard Biotools |
| 155Gd | CD57 | HCD57 | Standard Biotools |
| 156Gd | CD183 (CXCR3) | G025H7 | Standard Biotools |
| 158Gd | CD185 (CXCR5) | J252D4 | Standard Biotools |
| 159Tb | CD23 | EBVCS-5 | Biolegend (custom conjugated) |
| 160Gd | CD28 | CD28.2 | Standard Biotools |
| 161Dy | CD38 | HB-7 | Standard Biotools |
| 162Dy | CD49d | 9F10 | Biolegend (custom conjugated) |
| 163Dy | CD56 (NCAM) | NCAM16.2 | Standard Biotools |
| 164Dy | TCRγδ | B1 | Standard Biotools |
| 166Er | CD294 (CRTH2) | BM16 | Standard Biotools |
| 167Er | CD197 (CCR7) | G043H7 | Standard Biotools |
| 168Er | CD14 | 63D3 | Standard Biotools |
| 170Er | CD3 | UCHT1 | Standard Biotools |
| 171Yb | CD20 | 2H7 | Standard Biotools |
| 172Yb | CD66b | G10F5 | Standard Biotools |
| 173Yb | HLA-DR | LN3 | Standard Biotools |
| 174Yb | IgD | IA6-2 | Standard Biotools |
| 176Yb | CD127 (IL-7Ra) | A019D5 | Standard Biotools |
| 209Bi | CD11b | ICRF44 | Standard Biotools |

**Supplementary Figure E1: Comparative analysis of percentage of Th2 and T-reg cells in chronic rhinosinusitis (CRS).** Comparison of percentage (y-axis) of (A) Th2 and (B) T-reg cells in disease groups (controls, blue, *n*=6; CRS without polyps (CRSsNP), grey, *n*=6; CRS with nasal polyps (CRSwNP), green, n=4; and non-steroidal anti-inflammatory drugs (NSAIDs)-exacerbated respiratory disease (N-ERD, red, *n*=6). Box plots show the range with medians as horizontal lines and whiskers indicating minimum and maximum values. Groups showing no significant differences using the Kruskal-Wallis test followed by Dunn’s test (*p*>0.05).

**Supplementary Figure E2:** **Th2a cell percentages in chronic rhinosinusitis (CRS) disease groups stratified by allergy status.** Control allergic (Control-A, light blue, *n*=3), control non-allergic (Control-NA, blue, *n*=3), CRS without nasal polyps (CRSsNP) allergic (CRSsNP-A, light grey, *n*=3), CRSsNP non-allergic (CRSsNP-NA, grey, *n*=3), CRS with nasal polyps (CRSwNP) allergic (CRSwNP-A, green, *n*=1), CRSwNP non-allergic (CRSwNP-NA, light green, *n*=3), non-steroidal anti-inflammatory drugs (NSAIDs)-exacerbated respiratory disease (N-ERD)-allergic (N-ERD-A, red, n=3) and N-ERD non-allergic (N-ERD-NA, orange, *n*=3) groups. Box plots show the range with medians as horizontal lines and whiskers indicating minimum and maximum values. Groups showing no significant differences using the Kruskal-Wallis test followed by Dunn’s test (*p*>0.05).

**Supplementary Figure E3: Proportions of B cell sub-types in allergy-atratified chronic rhinosinusitis (CRS) disease groups.** Comparison of median percentage (y-axis) B cell subtypes sub-clustered via FlowSOM and identified as B1-B10 in control allergic (Control-A, light blue, *n*=3), control non-allergic (Control-NA, blue, *n*=3),CRS without nasal polyps (CRSsNP) allergic (CRSsNP-A, light grey, *n*=3), CRSsNP non-allergic (CRSsNP-NA, grey, *n*=3), CRS with nasal polyps (CRSwNP) allergic (CRSwNP-A, green, *n*=1), CRSwNP non-allergic (CRSwNP-NA, light green, *n*=3), non-steroidal anti-inflammatory drugs (NSAIDs)-exacerbated respiratory disease (N-ERD)-allergic (N-ERD-A, red, *n*=3) and N-ERD non-allergic (N-ERD-NA, orange, *n*=3) groups. Box plots show the range with medians as horizontal lines and whiskers indicating minimum and maximum values. Stars represent statistically significant differences between groups after Kruskal-Wallis test followed by Dunn’s test (*: *p*≤0.05).

**Supplementary Figure E4: Median metal intensity (MMI) of CD45RA, CXCR5 and CD49d in B cell subtypes in allergic and non-allergic patients**. (A-C) MMI of (A) CD45RA, (B) CXCR5, and (C) CD49d in B cell sub-clusters B1-B5 (left panels) and B 6 to B10 (right panels) in allergic (*n*=10) and non-allergic individuals (*n*=12). Stars represent statistically significant differences between groups using the Mann-Whitney-U Test (*: *p*≤0.05).

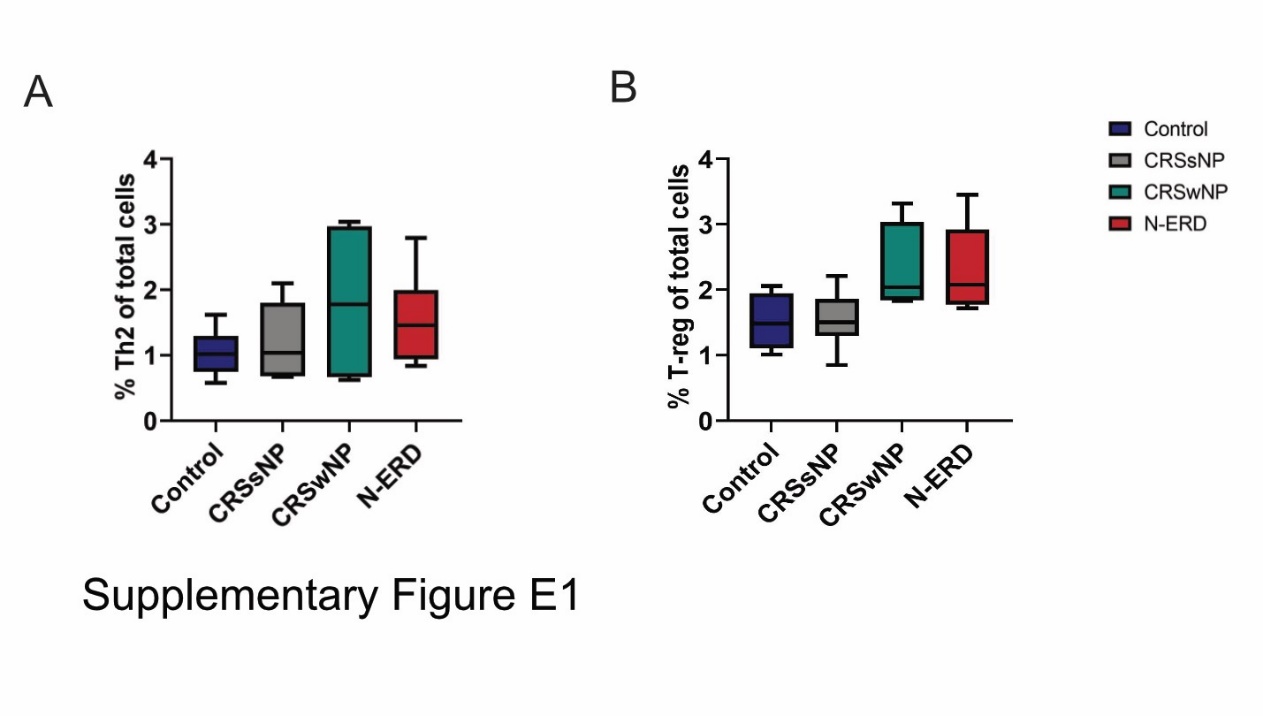

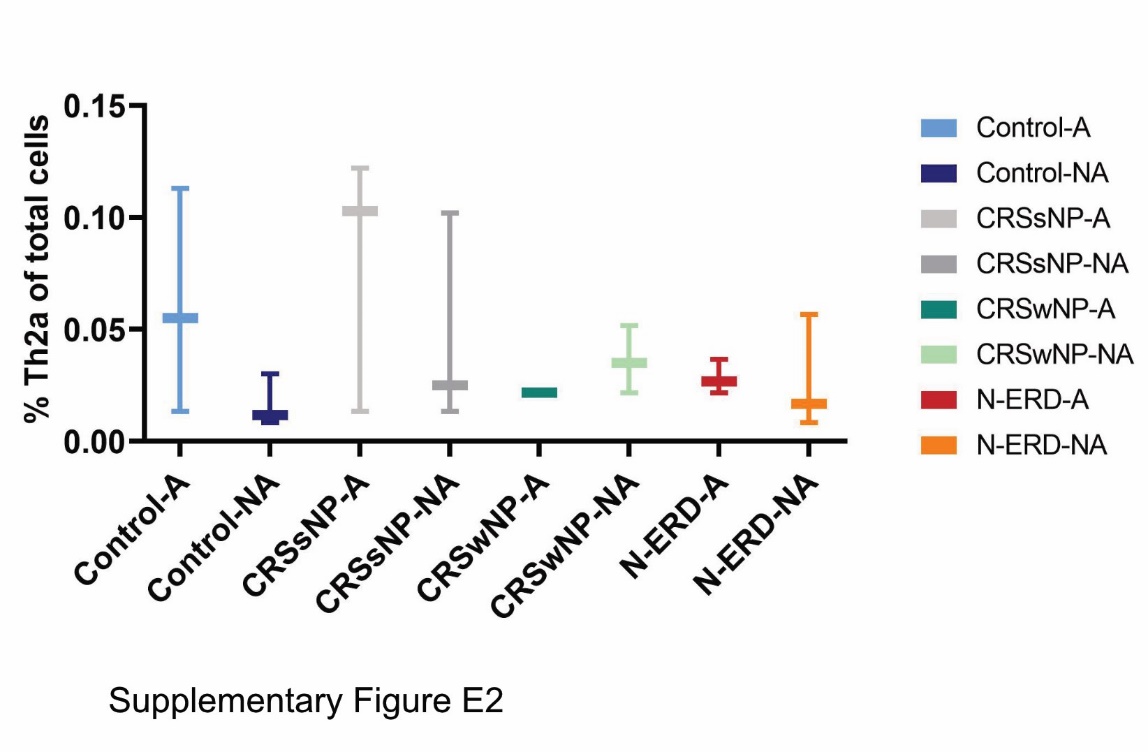

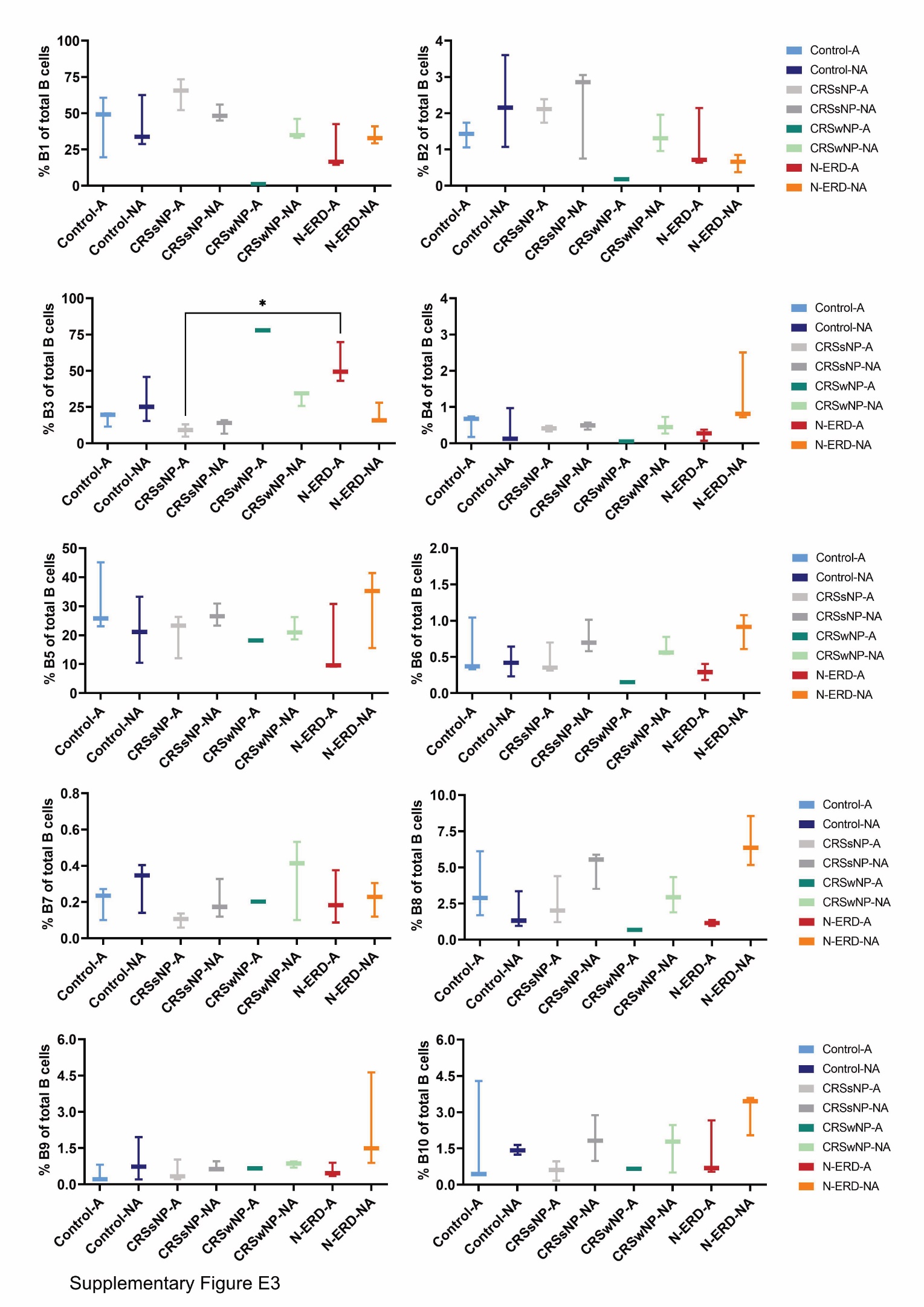

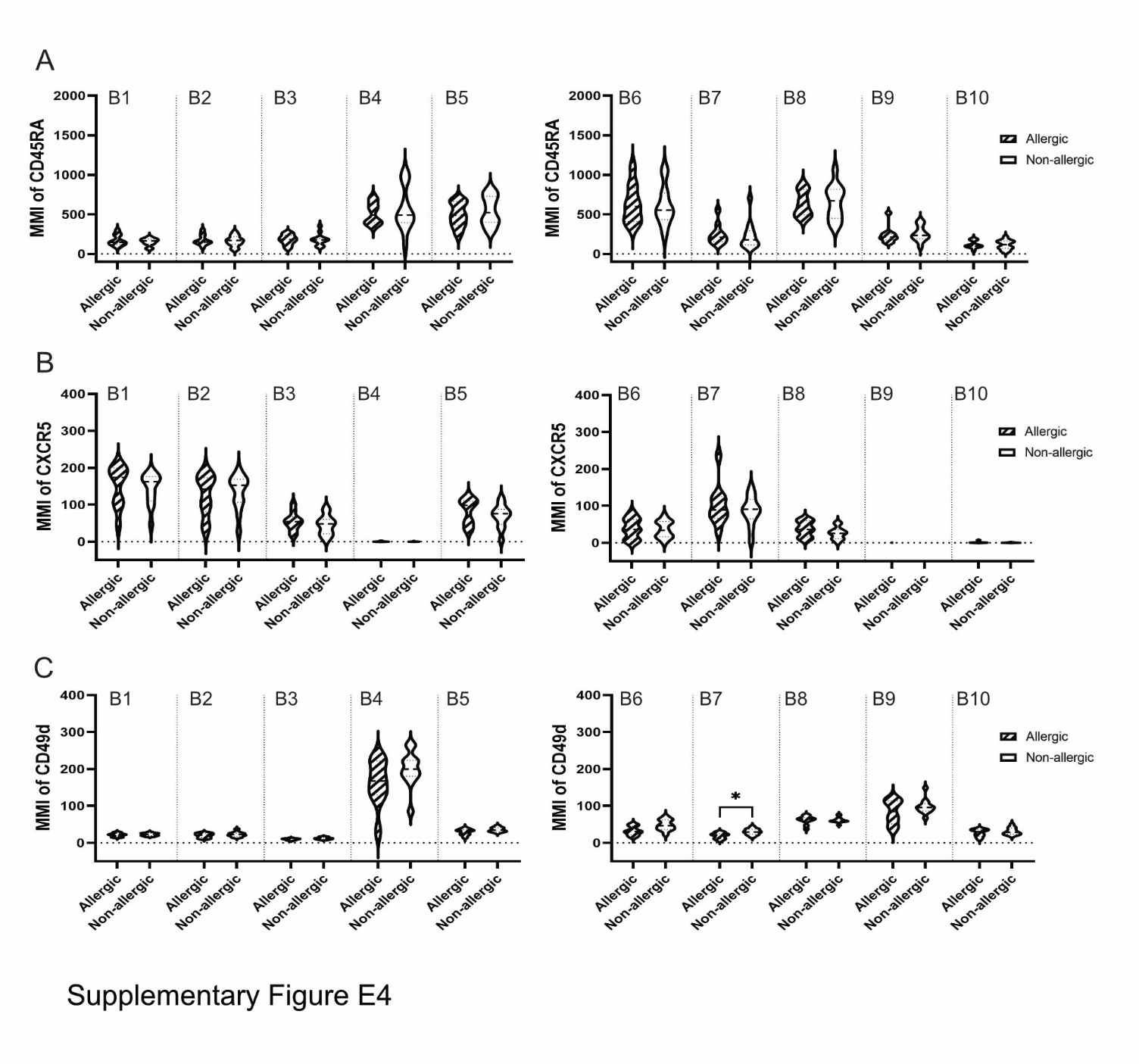

3. **REFERENCES**

1.	Rocha-Hasler, M., Muller, L., Wagner, A., Tu, A., Stanek, V., Campion, N.J., Bartosik, T., Zghaebi, M., Stoshikj, S., Gompelmann, D., et al. (2022). Using mass cytometry for the analysis of samples of the human airways. Front Immunol *13*, 1004583. 10.3389/fimmu.2022.1004583.

2.	Bagwell, C.B., Inokuma, M., Hunsberger, B., Herbert, D., Bray, C., Hill, B., Stelzer, G., Li, S., Kollipara, A., Ornatsky, O., and Baranov, V. (2020). Automated Data Cleanup for Mass Cytometry. Cytometry A *97*, 184-198. 10.1002/cyto.a.23926.
